# Statistical Analysis Plan for the Dig-RHD trial evaluating Digoxin in patients with rheumatic heart disease

**DOI:** 10.64898/2025.12.20.25342732

**Authors:** Aman Rastogi, Arpita Ghosh, Niveditha Devasenapathy, Shyamashree Biswas, Ganesan Karthikeyan

## Abstract

This Statistical Analysis Plan (SAP) describes the planned analyses for the Dig-RHD trial, a multicenter, randomized, double-blind, placebo-controlled study evaluating the efficacy and safety of digoxin in adults with rheumatic heart disease (RHD) and heart failure or atrial fibrillation/flutter. The primary objective is to assess the effect of digoxin, compared with placebo, on the composite endpoint of all-cause mortality or new or worsening HF over a follow-up period of up to 36 months. Secondary endpoints include time to death due to any cause, time to HF-related death, time to first and recurrent hospitalizations for HF, time to new-onset or worsening HF, and changes in quality of life (EQ-5D-5L scores). Efficacy analyses will be conducted in the modified intention-to-treat (mITT) population, defined as all randomized participants who received at least one dose of study medication. Time-to-event outcomes will be analyzed using Cox proportional hazards models, with additional sensitivity analyses including restricted mean survival time and joint frailty models for recurrent events. Quality of life outcomes will be analyzed using mixed-effects models for repeated measures. All analyses will be performed using R statistical software. This SAP was finalized prior to database lock and unblinding, in accordance with ICH-E9 guidelines, to ensure transparency and methodological rigor.

## 1. Administrative information

### 1.1 Study identifiers

- Protocol Version 1.3 16 June 2022
- Clinical trial registration: CTRI/2021/04/032858

### 1.2 Revision history

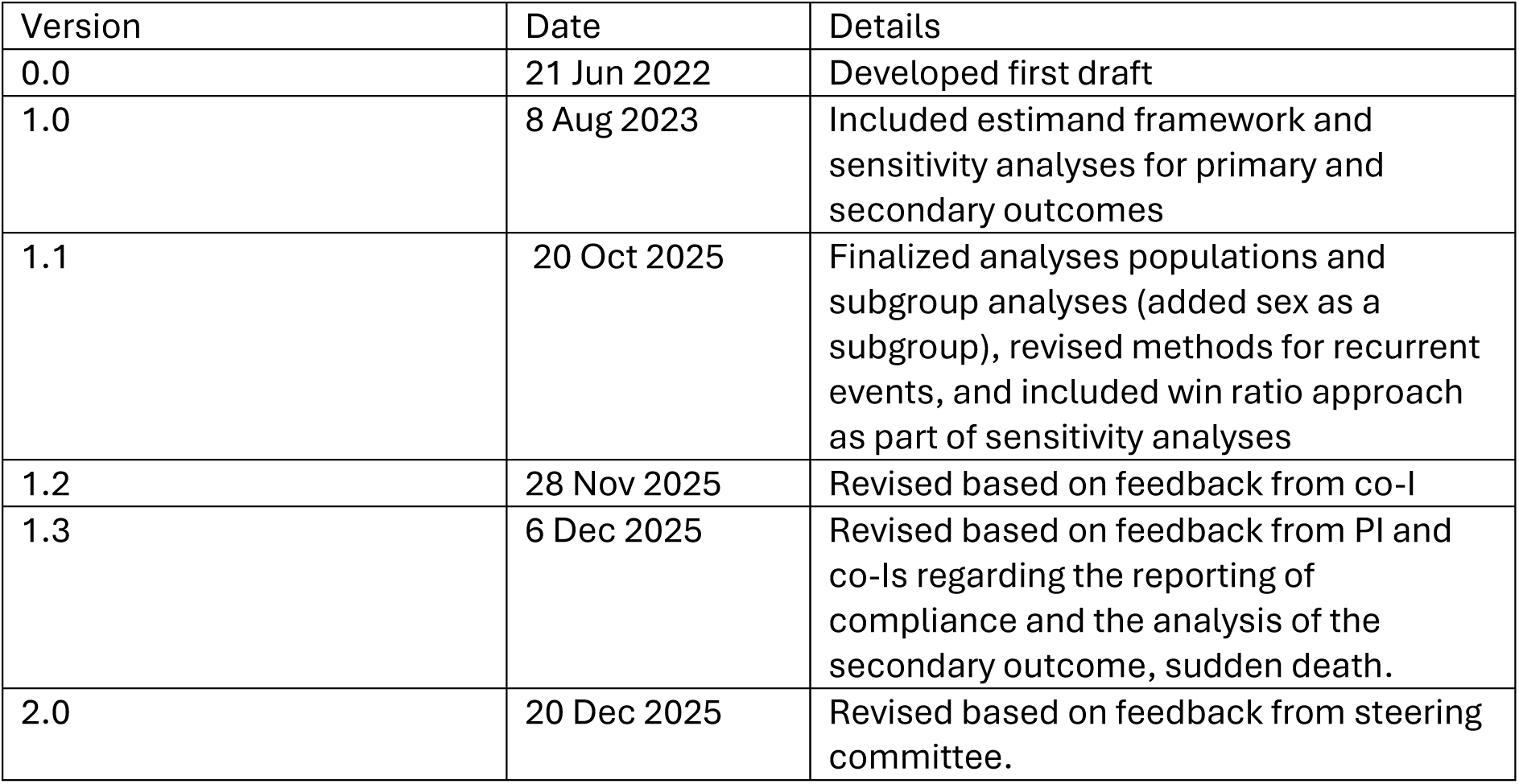

| Version | Date | Details |
| --- | --- | --- |
| 0.0 | 21 Jun 2022 | Developed first draft |
| 1.0 | 8 Aug 2023 | Included estimand framework and sensitivity analyses for primary and secondary outcomes |
| 1.1 | 20 Oct 2025 | Finalized analyses populations and subgroup analyses (added sex as a subgroup), revised methods for recurrent events, and included win ratio approach as part of sensitivity analyses |
| 1.2 | 28 Nov 2025 | Revised based on feedback from co-I |
| 1.3 | 6 Dec 2025 | Revised based on feedback from PI and co-Is regarding the reporting of compliance and the analysis of the secondary outcome, sudden death. |
| 2.0 | 20 Dec 2025 | Revised based on feedback from steering committee. |

### 1.3 Contributors to the statistical analysis plan

| Name | Role on study | Contribution to the SAP |
| --- | --- | --- |
| Aman Rastogi | Study statistician | Developed the SAP |
| Arpita Ghosh | Senior statistician (Co-I) | Reviewed and edited every version |
| Niveditha Devasenapathy | Co-I | Helped develop first draft and reviewed and provided critical input to all versions |
| Shyamashree Biswas | Research Consultant | Reviewed and provided critical inputs to all versions |
| Ganesan Karthikeyan | PI | Reviewed, provided critical input, and approved final version |

### 1.4 Approvals

The undersigned have reviewed this plan and approve it as final. They find it to be consistent with the requirements of the protocol as it applies to their respective areas. They also find it to be compliant with ICH-E9 principles and particularly confirm that this analysis plan was developed in a completely blinded manner, i.e. without knowledge of the effect of the intervention(s) being assessed.

| Signature | Date |
| --- | --- |
| Aman Rastogi<br>*aman* | 20 Dec 2025 |
| Arpita Ghosh<br>*Arpita Ghosh* | 20 Dec 2025 |
| Niveditha Devasenapathy<br>*Niveditha* | 20th December 2025 |
| Shyamashree Biswas<br>*sbiswas* | 20th December 2025 |
| Ganesan Karthikeyan<br>*GK* | 20th December 2025 |

## 2. Introduction

### 2.1 Background

Digoxin is widely used in the medical management of patients with rheumatic heart disease (RHD), for rate control in those with atrial fibrillation or atrial flutter (AF), and for treatment of heart failure (HF). Observational data from older patients with non-valvular AF suggest higher risk of mortality among those who received digoxin. However, patients with RHD are younger, without associated comorbidity, and it is unclear if digoxin will have similar adverse effects in these patients.

Extrapolating the results of studies in patients without valve disease, to patients with RHD may result in underuse of an inexpensive treatment which has proven benefit in reducing hospitalization for worsening HF. There is therefore a clear need for an adequately powered randomized controlled trial to evaluate the efficacy and safety of digoxin in this population.

### 2.2 Objective

The primary objective is to evaluate the effect of digoxin therapy compared to placebo (on a background of standard care) on the composite of all-cause death and new or worsening HF at 2 years. The secondary objectives are to determine the effect of digoxin on all cause death, death due to HF, sudden death, hospitalisation due to HF, new-onset or worsening HF and quality of life at 2 years [1].

## 3. Study Methods

### 3.1 Trial design

Two-arm, 1:1 randomised, multi-centre, placebo-controlled, parallel-group, superiority trial.

#### 3.1.1 PICOT

**P**: Adult patients with echocardiographically confirmed RHD with HF or AF

**I**: Digoxin 0.25 mg or 0.125 mg once daily with up to 2 “digoxin-free” days per week, PLUS standard evidence-based HF medications

**C**: Matching placebo PLUS standard evidence-based HF medications

**O**: A composite of all-cause death and new or worsening HF

**T**: Till 36 months after randomization

#### 3.1.2 Study Design

Two-arm, 1:1 randomised, multi-centre, placebo-controlled, parallel-group, superiority trial.

#### 3.1.3 Trial population

##### 3.1.3.1 Inclusion criteria

- Age ≥ 18 years
- RHD documented by echocardiography
- Already on treatment with digoxin as advised by their treating doctors [or]
- Not already on digoxin, but have either of the following conditions:

`o` HF - diagnosed in the presence of current or past clinical symptoms (limitation of activity, fatigue, and dyspnea, orthopnea or paroxysmal nocturnal dyspnea), signs (edema, elevated jugular venous pressure, or rales), or radiologic evidence of pulmonary congestion
`o` Atrial fibrillation or flutter - documented on a 12-lead ECG

##### 3.1.3.2 Exclusion criteria

- Significant renal insufficiency (serum creatinine >2 mg/dL)
- Uncorrected hypokalemia (<3.2 mmol/L) or hyperkalemia (>5.5 mmol/L)
- Presence of sinus bradycardia (HR <50/min), II- or III-degree AV block, sick sinus syndrome or pre-excitation
- Physicians do not wish to start or withhold digoxin
- Coexisting coronary artery disease or other cardiac conditions (hypertrophic, dilated or restrictive cardiomyopathies)
- Patients with mechanical heart valves (however, patients who undergo mechanical valve replacement surgery during the course of the study may be continued on study drug as required)
- Pregnant women
- Presence of severe comorbid conditions which may limit life expectancy to <2 years

### Trial drug initiation and follow-ups

Once randomized, the study medication will be initiated at the next scheduled dose. In patients already on digoxin, the study medication will replace open-label digoxin at the same dose as prescribed before. The study medication will be supplied to the trial participant during their hospital visits. Each kit consists of hundred tablets of 0.25 mgs. After randomization, patients will be seen every 6 months by their treating physicians for a minimum duration of 18 months or until the end of trial whichever is the earliest and a maximum of 36 months. Participants are encouraged to follow up within a window period of +/- 3 months from the scheduled date of visit. However, if the patient visits beyond this window period, then the visit details such as medication compliance and concomitant medications are being captured to the visit closest to the planned visit [1].

### Timing of outcome assessments

Information about all events (death, HF, hospitalisation) will be collected during the follow-up visits at 6, 12, 18, 24, 30 and 36 months or during telephonic reminder call at 3-month intervals or unscheduled visits to the hospital. Self-reported QoL will be collected only at baseline,12-, 18-, and 24-month visits.

### 3.2 Randomization

The randomization process is described in full within the clinical trial protocol [1]. In brief, participants were randomised in a 1:1 ratio to the treatment groups using randomly permuted blocks of varying sizes. The allocation sequence was stratified by hospital sites and the presence or absence of AF at enrolment through an Interactive Web Response System (IWRS) integrated with the Electronic Data Capture (EDC) system.

### 3.3 Sample size

Patients with an indication for digoxin would have a 20% risk of death at 2 years [2, 3]. In addition, the risk of hospitalization for new or worsening HF at 2 years would be about 10% in these patients. We, therefore, expect that the primary composite consisting of all-cause death and new or worsening HF, will occur in about 30% of patients at 2 years. Allowing for a 5% loss to follow-up per year, with 1800 participants, we will be able to detect a 20% relative reduction in the primary composite outcome with at least 83% power at a two-sided α of 0.05.

### 3.4 Statistical interim analyses and stopping guidance

Efficacy outcomes were not analyzed during interim analyses, and no statistical stopping rules were applied. The trial stopping criteria were solely based on safety concerns, poor recruitment, or poor data quality. One interim review of the overall data was performed in October 2024 after 256 primary composite events had occurred. Number of deaths, discontinuations attributed to suspected digoxin toxicity and HF events in the overall trial population were presented to the independent Data Safety and Monitoring Board (DSMB). These numbers were assessed against numbers expected in routine practice. Subsequently, in a closed session, group-blinded results were shared with the DSMB, which recommended continuing the trial without any modifications.

### 3.5 Timing of final analysis

Final analysis will be performed when last patient’s last follow-up visit data have been entered, blinded review of data has been conducted for the final quality check, and SAP has been published. SAP will be made available in the public domain before breaking the blind.

## 4. Statistical principles

### 4.1 Confidence intervals and p-values

All tests will be two-sided with a nominal level of 𝛼 set at 5%. No multiplicity adjustment will be made as there is only one primary efficacy outcome.

The secondary efficacy outcomes measure different aspects of the same underlying treatment effect that is being evaluated in the primary efficacy outcome. Therefore, the secondary outcomes provide only supporting evidence. Hence, no type I error adjustment is proposed to account for multiplicity of secondary efficacy outcomes.

### 4.2 Compliance to trial medication and protocol deviations

**Compliance:** During hospital visits, patients were assessed by study coordinator for compliance based on pill count and interview. Patients were categorized as being compliant for a period if they took at least 80% of the prescribed medication for that period, in the absence of any contraindication. Following summaries will be given by treatment arm:

1. Number and proportion of patients who are compliant at each visit.
2. Number and proportion of patients who temporarily discontinue the trial drug during at least one visit; number of temporary discontinuations and their reasons (subject to availability of reasons).
3. Number and proportion of patients who permanently discontinue the trial drug and their reason for discontinuation (subject to availability of reasons).

**Protocol deviation:** Following two protocol deviations will be summarized:

1. Number of randomized patients who are ineligible, by arm.
2. All included randomized patients who permanently discontinue their medication without any medical advice by physician. This will be reported by arm and time of discontinuation.

### 4.3. Analysis populations

#### 4.3.1 Intention-to-treat (ITT)

The ITT population is defined as all **patients who were randomized**.

#### 4.3.2 Modified intention-to-treat (mITT)

The mITT population is defined as all randomized patients who have taken **at least one dose of assigned treatment**.

Baseline characteristics of study participants and protocol deviations will be reported for the ITT population. Medication adherence, concomitant medications and SAEs will be reported for the mITT population. Primary and secondary outcomes will be analysed using the mITT population.

### 4.4 Subject disposition and baseline patient characteristics

#### 4.4.1 Subject disposition

The flow of participants through the trial will be displayed in a CONSORT (Consolidated Standards of Reporting Trials) diagram. It will include the number enrolled and randomised, the number receiving their allocated treatment, the number of deaths, withdrawals, and lost to follow-up, and the number randomized and included in the primary analysis (mITT) with reasons for non-inclusion for the primary outcome analysis. In addition, we will provide an enrolment summary by centre: the number of participants recruited, the number of days to recruit, and the number of participants recruited in each stratum.

#### 4.4.2 Baseline characteristics

Baseline characteristics of the mITT population – age, sex, education, employment status, BMI, blood pressure, heart rate, medical history, medications, whether on Digoxin or not, NYHA class, rhythm status, laboratory parameters and quality of life score – will be summarized by treatment groups. For categorical variables, the number and percentage of participants within each category (with a category for missing data) will be presented. Percentages will be calculated according to the number of patients for whom data are available. Continuous data will be summarised by mean, standard deviation, median, interquartile interval (Q1-Q3), range, and the number of missing observations, if any. No statistical test will be performed on baseline characteristics.

## 5. Analysis

### 5.1 Outcome definitions

#### 5.1.1 Primary outcome

**The primary outcome is a composite of all-cause death or new or worsening HF,** defined as the time from randomization to the first occurrence of any component of the composite till 36 months or until the end of the trial. New onset HF is diagnosed if any 2 of the 3 following criteria are present: (i) symptoms (dyspnea on exertion or at rest, orthopnea, nocturnal paroxysmal dyspnea, or ankle edema) or signs (rales, increased jugular venous pressure or ankle edema) of congestive HF, (ii) radiologic signs of pulmonary congestion, and (iii) treatment with diuretics. Criteria for worsening of HF is either (i) escalation of oral diuretics (an increase in diuretic dose or the addition of another class of diuretics), or (ii) need for intravenous diuretics in an outpatient or emergency department, or (iii) hospital admission for treatment of HF, all with or without deterioration by at least one NYHA class.

#### 5.1.2 Secondary outcomes

1. **Time to death due to any cause** will be defined as the time from randomization to the occurrence of death due to any cause till end of study or 36 months.
2. Quality of life (EQ-5D-5L) scores at 12, 18 and 24 months
3. **Time to first occurrence of the composite of HF-related death or new-onset or worsening HF** will be defined as the time from randomization to the occurrence of HF related death or new or worsening HF, whichever occurred earlier, till end of study or 36 months.
4. **Time to first occurrence of the composite of HF-related death or hospitalisation for HF** will be defined as the time from randomization to the occurrence of HF related death or new or worsening HF, whichever occurred earlier, till end of study or 36 months.
5. **Time to first HF** will be defined as the time from randomization to the occurrence of new or worsening HF till end of study or 36 months.
6. **Time to HF (all events)** will be defined as the time from randomization to each HF till end of study or 36 months.
7. **Time to first hospitalisation for HF** will be defined as the time from randomization to the first hospitalization for HF till end of study or 36 months.
8. **Time to hospitalisation for HF (all events)** will be defined as the time from randomization to each hospitalisation for HF till end of study or 36 months.
9. **Time to death due to HF** will be defined as the time from randomization to occurrence of death due to HF till end of study or 36 months.
10. **Time to sudden death,** defined as death occurring within 24 hours of symptom-onset or worsening

#### 5.1.3 Safety outcomes

Serious adverse events (SAEs):

1. Any hospitalization unrelated to valve disease
2. Clinically suspected digoxin toxicity leading to discontinuation of trial medication with or without hospitalization

### 5.3 Analysis methods

#### 5.3.1 Analysis of primary outcome

##### 5.3.1.5 Estimand

The estimand for the primary outcome is defined by the following components:

- **Target population**: mITT population, defined as all randomized participants who received at least one dose of study medication. This aligns with a principal stratum strategy for participants who initiated randomized treatment, assuming that under double-blind conditions the decision to initiate treatment is not influenced by knowledge of allocation.
- **Intervention**: addition of digoxin as treatment strategy over and above the standard treatment for HF. Hence dose and dosage changes along with decision to temporarily or permanently discontinue by the physician is part of the treatment strategy.
- **Variable (endpoint)**: time from randomization to the first occurrence of the composite of death from any cause, or new or worsening HF.
- **Summary measure**: hazard ratio (HR) from a mixed-effects Cox proportional hazards model
- **Intercurrent events and strategies**

| Intercurrent event | Strategy |
| --- | --- |
| Failure to initiate randomized treatment | Principal stratum (participants who initiated treatment) |
| Discontinuation of digoxin due to suspected toxicity or relief of symptoms following percutaneous/valve procedures | Treatment policy |
| Switch to open-label digoxin | Treatment policy |
| Initiation of diuretics | Composite variable |

##### 5.3.1.2 Main analysis

The endpoint is a composite of all-cause death or new or worsening HF, defined as the time from randomization to the first occurrence of any component of the composite till 36 months or until the end of the trial. Participants who did not experience any component event during the study observation period, who withdrew consent, or who were lost to follow-up will be censored at the date of their last contact. Participants with no post-randomization follow-up will be censored on the date of randomization.

The outcome will be summarized using Kaplan–Meier plots showing cumulative event rates by treatment arm with 95% confidence intervals (CIs), and the number of participants at risk will be displayed along the x-axis. A Cox proportional hazards model will be used to estimate the effect of treatment, including fixed effects for treatment group and baseline rhythm status (presence vs. absence of atrial fibrillation or atrial flutter) and a random effect for hospital site to account for clustering of participants within sites [4]. The treatment effect will be reported as the adjusted HR with its 95% CI. Potential non-proportional hazards will be explored in sensitivity analyses.

##### 5.3.1.3 Adjusted analysis

No other variables will be adjusted for; however, if any adjusted analyses are performed, they will be conducted post hoc and considered exploratory in nature.

##### 5.3.1.4 Subgroup analysis

Sub-group analysis will be conducted for the primary outcome for following subgroups (sex was added after protocol was published), irrespective of whether there is a statistically significant main treatment effect:

1. AF at baseline (yes vs. no)
2. Digoxin therapy at the time of enrolment (yes vs. no)
3. Sex (male or female)

The model used in the main analysis will be applied, with the addition of the main effect for the subgroup (if not already included) and a treatment-by-subgroup interaction term. Heterogeneity of treatment effect across subgroups levels will be assessed by testing the statistical significance of the interaction term. For each level of the subgroup, HRs with corresponding 95% CIs will be presented on a forest plot, along with the p-value for interaction.

Any other subgroup analyses will be considered post-hoc and exploratory.

##### 5.3.1.5 Sensitivity analysis

The following sensitivity analyses will be performed to explore the robustness of the findings from the main analysis and to improve interpretability of. These analyses will be regarded as exploratory.

1. The proportional hazards assumption will be checked visually, and the primary composite outcome will be analysed using restricted mean survival time (RMST) [5–8]. Complementary log-log survival will be plotted against log(time) for treatment groups and baseline AF status. Parallel curves support proportional hazards. Scaled Schoenfeld residuals for each covariate (treatment, baseline AF) will be plotted against event time, with a smoothed (e.g., loess) curve to assess potential time trends. With proportional hazards, the loess curve should be parallel to the x-axis. RMST up to 36 months (or the last common follow-up time, if earlier) will be estimated as the area under the Kaplan–Meier survival curve for each treatment arm, representing the average event-free survival time within that period. The treatment effect will be expressed as the difference in RMST (treatment − control), corresponding to the average delay (or gain) in occurrence of the composite event associated with treatment. The associated two-sided 95% CI and p-value will be reported [9, 10]. This approach does not rely on the proportional hazards assumption.
2. An unmatched win-ratio analysis will be performed. Win ratio method places the components of the composite into a hierarchy from most to least clinically important, thereby giving greater priority to the more important ones. Each participant in the treatment arm will be compared with all participants in the control arm, forming all possible pairs (unmatched). For each pair, outcomes will be evaluated hierarchically: (1) all-cause death, (2) number of heart-failure (HF) events, and (3) time to first HF event. The win-ratio, its 95% CI, and p-value will be reported along with the proportion of ties [11, 12]. In addition, the win difference that takes the number of tied comparisons into account will be reported [12, 13].
3. If death of the participant occurs within 3 days after a recorded HF, this will be counted as a single event and will be considered as a death for the primary outcome analysis, and date of event will be the date of death.
4. The assumption of non-informative censoring will be assessed by examining reason for censoring and characteristics of censored patients in the two treatment arms.

#### 5.3.2 Analyses of secondary outcomes

1. Time to death due to any cause.

Time to death due to any cause will be analysed using the same approach as described in the main analysis of primary outcome, i.e. using Kaplan-Meier plots and Cox proportional hazards model with fixed effects for treatment and baseline AF status (presence vs absence) and a random effect for hospital site. This outcome will be additionally analysed after relaxing the proportional hazards assumption using RMST and we will report the RMST difference along with 95% CI.

2. Time to first new onset or worsening HF
3. Time to first hospitalisation for HF

For both the time-to-event outcomes, death due to any cause will be treated as competing event. As part of descriptive analysis, cumulative incidence curves showing the cumulative probability of event over time will be presented. The effect of treatment will be estimated using Cox proportional hazards model and the cause-specific HR will be reported [16]. As before, the model will include fixed effects for treatment and baseline AF status and a random effect for hospital sites.

The distribution of competing events – frequency and distribution across treatment arms, will be examined. As sensitivity analysis, Fine & Gray model will be fit and subdistribution HR will be reported. In addition, a multiple imputation approach that extends the Cox model for missing follow-up will be explored [17].

4. Time to first occurrence of the composite of HF-related death or new-onset or worsening HF.
5. Time to first occurrence of the composite of HF-related death or hospitalisation for HF.
6. Time to occurrence of death due to HF.

These outcomes will be analysed in a similar manner - using Cox proportional hazards model. Non-HF related deaths will be treated as competing events for all three outcomes and patients dying due to non-HF related reasons will be censored. The treatment effect will be a cause-specific HR, reported with its 95% CI. If descriptive analyses of competing events suggest that sensitivity analyses for competing risks are needed, a multiple imputation approach recommended by Gregson at al. will be taken [16].

7. Time to HF (all events)
8. Time to hospitalisation for HF (all events)

Recurrent HFs and HF hospitalizations will be analysed using a joint frailty model [18–20] that simultaneously models the recurrent event process and the terminal event (death), while accounting for correlation of events within patients and for between-centre heterogeneity. The joint frailty model will assume proportional hazards for both the recurrent event intensity and for the hazard of death and will use a shared frailty structure linking the two processes so that informative censoring by death is accommodated. Both submodels will include fixed effects for treatment and baseline AF status and two random effects for the frailty parameters – 1) a centre-level frailty to capture hospital site effects, and 2) a patient-level frailty to capture unobserved patient heterogeneity and within-patient correlation of recurrent events. Both frailty terms will be assumed to follow a log-normal distribution [21]. The analysis will use gap time between events; that is, time to the first event will be measured from randomization to the first event, time to the second event will be measured from the first to the second event, and so on. Patients who do not die during follow-up will be censored at their last known contact date. HRs for recurrent event and death will be reported along with their 95% CIs [22, 23]. The estimated centre and patient frailty variances, and the association parameter will be reported with 95% CIs; these quantify between-centre heterogeneity, within-patient heterogeneity, and the dependence between recurrent events and death, respectively. The model will be estimated using the *frailtypack* package in R [23].

9. Quality of life (EQ-5D-5L) scores at 12, 18 and 24 months.

Domain wise responses at each time point will be displayed graphically. QoL scores at 12, 18 and 24 months will be analysed using mixed model linear regression for repeated measures (MMRM) [24, 25]. Fixed covariates will include the EQ-5D-5L score at baseline visit, treatment arm, visit (12-, 18- or 24-months), treatment arm by visit interaction and AF status at baseline. Correlation between patients from the same site will be modelled using random effects. Correlation between repeated measures on the same patient will be handled by specifying that residual errors are correlated and will be modelled using unstructured covariance. The effect of the intervention will be estimated as the adjusted mean difference in scores at 12, 18 and 24 months, along with their 95% CIs. This analysis will include all subjects with a baseline EQ-5D-5L score and at least one follow-up visit score and will give valid estimates in case of data missing at random. EQ-5D-5L value set will be derived following the approach and code provided in *Jyani et al.* [26]. VAS scores will be similarly anlaysed.

As sensitivity analysis, patient’s missing follow-up scores after death, till 24 months, will be imputed with 0 or lowest India-specific score (-0.923) to signify worst quality of life and MMRM will be performed.

10. Time to sudden death

This outcome will be analysed using Cox proportional hazards model with fixed effects for treatment and baseline AF status (presence vs absence) and a random effect for hospital site, censoring other causes of death at the time they occur. The treatment effect will be a cause-specific HR, reported along with the 95% CI.

No adjusted or subgroup analyses are planned for secondary outcomes.

#### 5.3.3 Analysis of safety outcomes

All safety analyses will be descriptive in nature. For the following SAEs the number of events and the number and percentage of participants with one or more events will be presented by arm

1. any hospitalization unrelated to valve disease
2. clinically suspected digoxin toxicity leading to discontinuation of trial medication with or without hospitalization

## Statistical software

All statistical analyses will be performed using R version 4.4.1 or higher.

## Data Availability

All data produced in the present study are available upon reasonable request to the authors

## Appendix

Proposed tables and figures

**Table 1.**
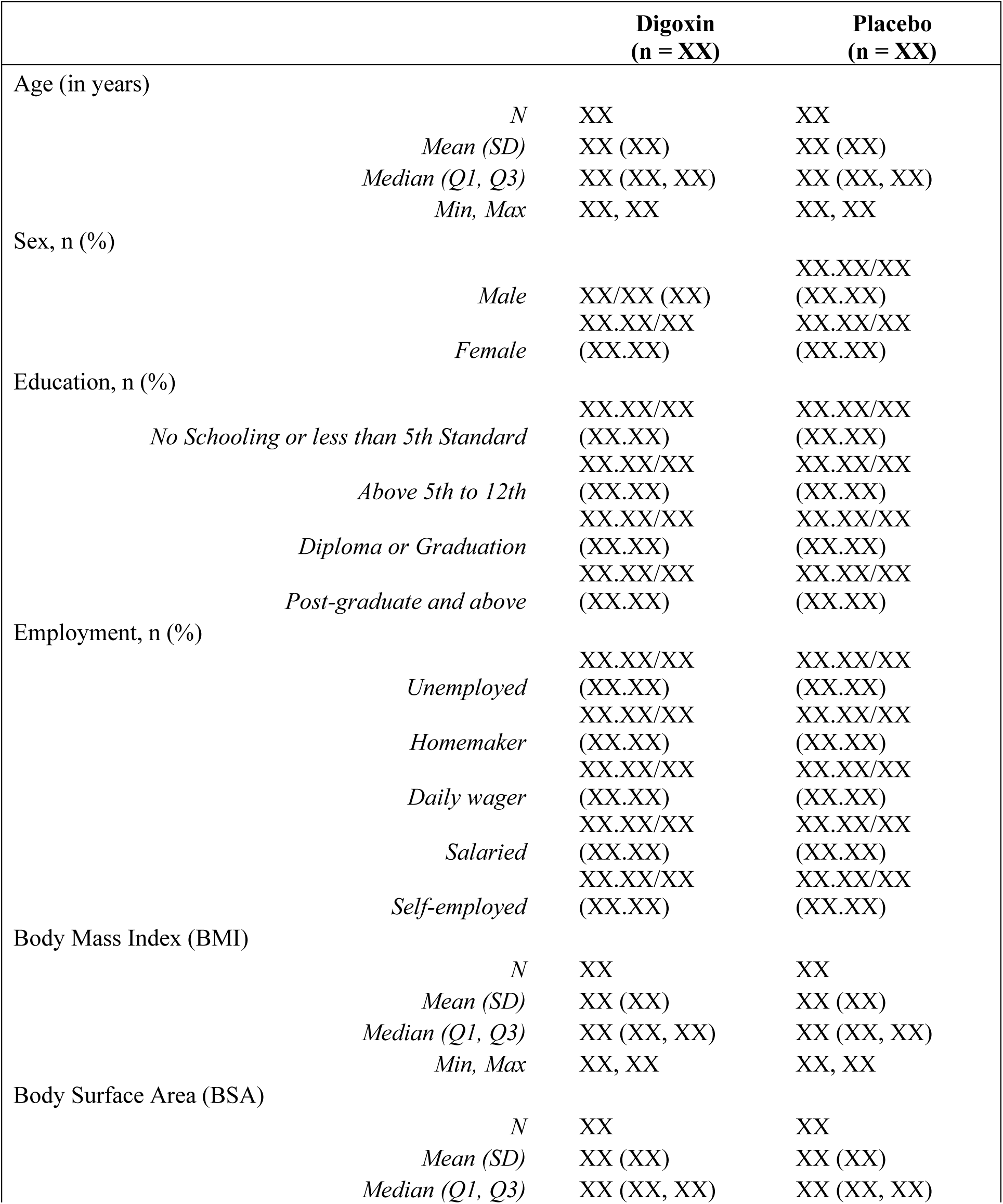
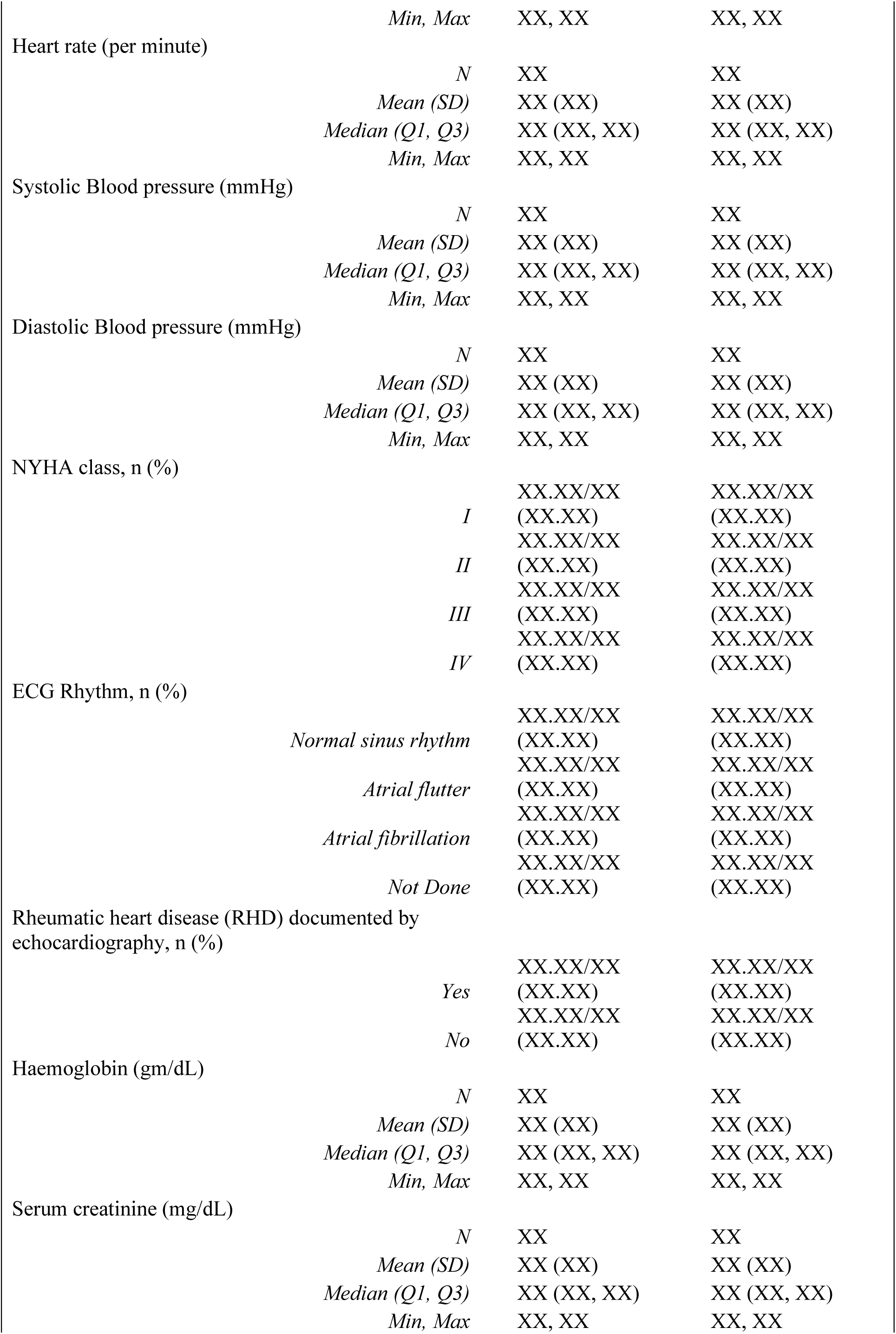
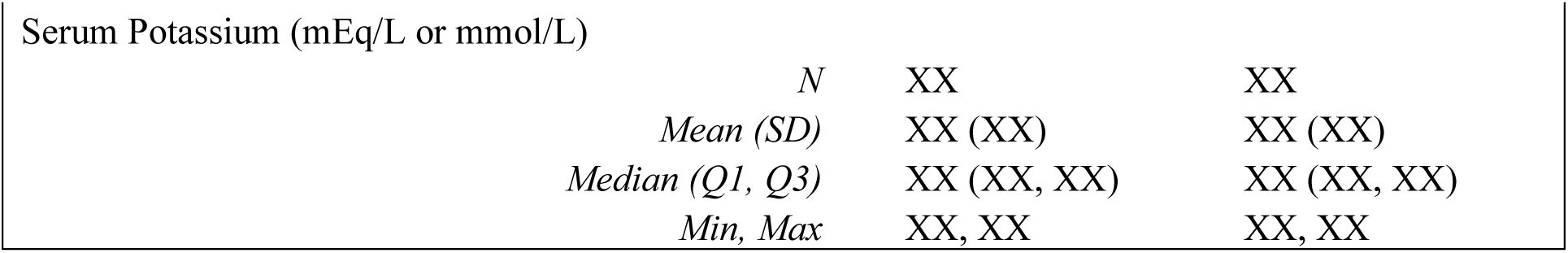
Baseline characteristics of Dig-RHD trial participants.

**Table 2.**
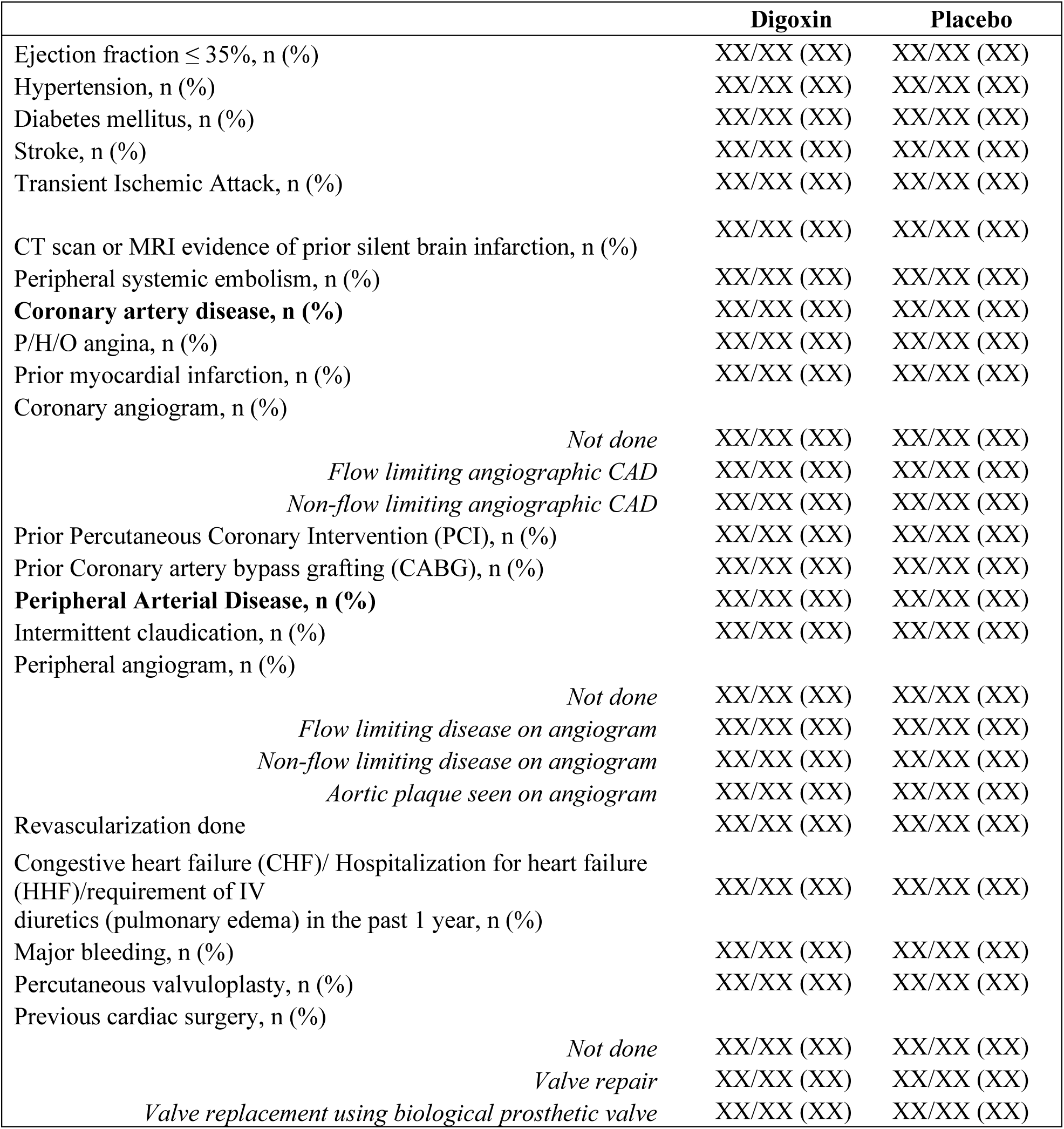
Past medical history.

**Table 3.** Echocardiography.

|  |  | Digoxin | Placebo |
| --- | --- | --- | --- |
| **Mitral valve** |  |  |  |
| Calcification, n (%) |  | XX/XX (XX) | XX/XX (XX) |
| Mitral stenosis, n (%) |  |  |  |
|  | No | XX/XX (XX) | XX/XX (XX) |
|  | Mild | XX/XX (XX) | XX/XX (XX) |
|  | Moderate | XX/XX (XX) | XX/XX (XX) |
|  | Severe | XX/XX (XX) | XX/XX (XX) |
| Mitral Valve Area (cm2) |  |  |  |
|  | *N* | XX | XX |
|  | *Mean (SD)* | XX (XX) | XX (XX) |
|  | *Median (Q1, Q3)* | XX (XX, XX) | XX (XX, XX) |
|  | *Min, Max* | XX, XX | XX, XX |
| Mitral regurgitation |  |  |  |
|  | No | XX/XX (XX) | XX/XX (XX) |
|  | Mild | XX/XX (XX) | XX/XX (XX) |
|  | Moderate | XX/XX (XX) | XX/XX (XX) |
|  | Severe | XX/XX (XX) | XX/XX (XX) |
| **Aortic valve** |  |  |  |
| Calcification, n (%) |  | XX/XX (XX) | XX/XX (XX) |
| Aortic stenosis, n (%) |  |  |  |
|  | No | XX/XX (XX) | XX/XX (XX) |
|  | Mild | XX/XX (XX) | XX/XX (XX) |
|  | Moderate | XX/XX (XX) | XX/XX (XX) |
|  | Severe | XX/XX (XX) | XX/XX (XX) |
| Jet velocity (cm/sec) |  |  |  |
|  | *N* | XX | XX |
|  | *Mean (SD)* | XX (XX) | XX (XX) |
|  | *Median (Q1, Q3)* | XX (XX, XX) | XX (XX, XX) |
|  | *Min, Max* | XX, XX | XX, XX |
| Peak gradient (mmHg) |  |  |  |
|  | *N* | XX | XX |
|  | *Mean (SD)* | XX (XX) | XX (XX) |
|  | *Median (Q1, Q3)* | XX (XX, XX) | XX (XX, XX) |
|  | *Min, Max* | XX, XX | XX, XX |
| Mean gradient (mmHg) |  |  |  |
|  | *N* | XX | XX |
|  | *Mean (SD)* | XX (XX) | XX (XX) |
|  | *Median (Q1, Q3)* | XX (XX, XX) | XX (XX, XX) |
|  | *Min, Max* | XX, XX | XX, XX |
| Aortic valve area (cm2) |  |  |  |
|  | *N* | XX | XX |
|  | *Mean (SD)* | XX (XX) | XX (XX) |
|  | *Median (Q1, Q3)* | XX (XX, XX) | XX (XX, XX) |
|  | *Min, Max* | XX, XX | XX, XX |
| Aortic regurgitation, n (%) |  |  |  |
|  | No | XX/XX (XX) | XX/XX (XX) |
|  | Mild | XX/XX (XX) | XX/XX (XX) |
|  | Moderate | XX/XX (XX) | XX/XX (XX) |
|  | Severe | XX/XX (XX) | XX/XX (XX) |
| Tricuspid valve |  |  |  |
| Calcification, n (%) |  | XX/XX (XX) | XX/XX (XX) |
| Tricuspid stenosis, n (%) |  | XX/XX (XX) | XX/XX (XX) |
| Mean diastolic gradient (mmHg) |  |  |  |
|  | *N* | XX | XX |
|  | *Mean (SD)* | XX (XX) | XX (XX) |
|  | *Median (Q1, Q3)* | XX (XX, XX) | XX (XX, XX) |
|  | *Min, Max* | XX, XX | XX, XX |
| Tricuspid regurgitation, n (%) |  |  |  |
|  | No | XX/XX (XX) | XX/XX (XX) |
|  | Mild | XX/XX (XX) | XX/XX (XX) |
|  | Moderate | XX/XX (XX) | XX/XX (XX) |
|  | Severe | XX/XX (XX) | XX/XX (XX) |
| Tricuspid valve annulus (mm) |  |  |  |
|  | *N* | XX | XX |
|  | *Mean (SD)* | XX (XX) | XX (XX) |
|  | *Median (Q1, Q3)* | XX (XX, XX) | XX (XX, XX) |
|  | *Min, Max* | XX, XX | XX, XX |
| **Pulmonary valve** |  |  |  |
| Calcification, n (%) |  | XX/XX (XX) | XX/XX (XX) |
| Pulmonary stenosis, n (%) |  | XX/XX (XX) | XX/XX (XX) |
| Pulmonary regurgitation, n (%) |  | XX/XX (XX) | XX/XX (XX) |
| Pulmonary hypertension, n (%) |  | XX/XX (XX) | XX/XX (XX) |
| Tricuspid regurgitation gradient (peak, mmHg) |  |  |  |
|  | *N* | XX | XX |
|  | *Mean (SD)* | XX (XX) | XX (XX) |
|  | *Median (Q1, Q3)* | XX (XX, XX) | XX (XX, XX) |
|  | *Min, Max* | XX, XX | XX, XX |
| Early Pulmonary regurgitation gradient (mmHg) |  |  |  |
|  | *N* | XX | XX |
|  | *Mean (SD)* | XX (XX) | XX (XX) |
|  | *Median (Q1, Q3)* | XX (XX, XX) | XX (XX, XX) |
|  | *Min, Max* | XX, XX | XX, XX |
| End Pulmonary regurgitation gradient (mmHg) |  |  |  |
|  | *N* | XX | XX |
|  | *Mean (SD)* | XX (XX) | XX (XX) |
|  | *Median (Q1, Q3)* | XX (XX, XX) | XX (XX, XX) |
|  | *Min, Max* | XX, XX | XX, XX |
| Pulmonary acceleration time (milli-second) |  |  |  |
|  | *N* | XX | XX |
|  | *Mean (SD)* | XX (XX) | XX (XX) |
|  | *Median (Q1, Q3)* | XX (XX, XX) | XX (XX, XX) |
|  | *Min, Max* | XX, XX | XX, XX |
| **Left ventricular (LV) dimension** |  |  |  |
| End systolic LV dimension (mm) |  |  |  |
|  | *N* | XX | XX |
|  | *Mean (SD)* | XX (XX) | XX (XX) |
|  | *Median (Q1, Q3)* | XX (XX, XX) | XX (XX, XX) |
|  | *Min, Max* | XX, XX | XX, XX |
| End diastolic LV dimension (mm) |  |  |  |
|  | *N* | XX | XX |
|  | *Mean (SD)* | XX (XX) | XX (XX) |
|  | *Median (Q1, Q3)* | XX (XX, XX) | XX (XX, XX) |
|  | *Min, Max* | XX, XX | XX, XX |
| Left atrium dimension (mm) |  |  |  |
|  | *N* | XX | XX |
|  | *Mean (SD)* | XX (XX) | XX (XX) |
|  | *Median (Q1, Q3)* | XX (XX, XX) | XX (XX, XX) |
|  | *Min, Max* | XX, XX | XX, XX |
| Aorta dimensions (mm) |  |  |  |
|  | *N* | XX | XX |
|  | *Mean (SD)* | XX (XX) | XX (XX) |
|  | *Median (Q1, Q3)* | XX (XX, XX) | XX (XX, XX) |
|  | *Min, Max* | XX, XX | XX, XX |
| LV ejection fraction (%) |  |  |  |
|  | *N* | XX | XX |
|  | *Mean (SD)* | XX (XX) | XX (XX) |
|  | *Median (Q1, Q3)* | XX (XX, XX) | XX (XX, XX) |
|  | *Min, Max* | XX, XX | XX, XX |
| Spontaneous Echo contrast, n (%) |  | XX/XX (XX) | XX/XX (XX) |
| Left atrium / Left Atrial Appendage thrombus, n (%) |  | XX/XX (XX) | XX/XX (XX) |
| Pericardial effusion, n (%) |  | XX/XX (XX) | XX/XX (XX) |

**Table 4.** Medication.

|  | Baseline | 6<sup>th</sup> month | 12<sup>th</sup> month | 18<sup>th</sup> month | 24<sup>th</sup> month | 30<sup>th</sup> month | 36<sup>th</sup> month |
| --- | --- | --- | --- | --- | --- | --- | --- |
| Dose prescribed, n (%) | n = XX | n = XX<sup>a</sup> | n = XX<sup>a</sup> | n = XX<sup>a</sup> | n = XX<sup>a</sup> | n = XX<sup>a</sup> | n = XX<sup>a</sup> |
| *0.25mg for 5 days* | XX/XX (%) | XX/XX (%) | XX/XX (%) | XX/XX (%) | XX/XX (%) | XX/XX (%) | XX/XX (%) |
| *0.25mg for 7 days* | XX/XX (%) | XX/XX (%) | XX/XX (%) | XX/XX (%) | XX/XX (%) | XX/XX (%) | XX/XX (%) |
| *0.125mg for 5 days* | XX/XX (%) | XX/XX (%) | XX/XX (%) | XX/XX (%) | XX/XX (%) | XX/XX (%) | XX/XX (%) |
| *0.125mg for 7 days* | XX/XX (%) | XX/XX (%) | XX/XX (%) | XX/XX (%) | XX/XX (%) | XX/XX (%) | XX/XX (%) |
| Trial medication status, n(%) |  | n = XX<sup>b</sup> | n = XX<sup>b</sup> | n = XX<sup>b</sup> | n = XX<sup>b</sup> | n = XX<sup>b</sup> | n = XX<sup>b</sup> |
| *Continuing* |  | XX/XX (%) | XX/XX (%) | XX/XX (%) | XX/XX (%) | XX/XX (%) | XX/XX (%) |
| *Temporary discontinuation* | NA | XX/XX (%) | XX/XX (%) | XX/XX (%) | XX/XX (%) | XX/XX (%) | XX/XX (%) |
| *Permanent discontinuation<sup>c</sup>* |  | XX/XX (%) | XX/XX (%) | XX/XX (%) | XX/XX (%) | XX/XX (%) | XX/XX (%) |
| Trial intervention compliance, n (%) |  | n = XX<sup>d</sup> | n = XX<sup>d</sup> | n = XX<sup>d</sup> | n = XX<sup>d</sup> | n = XX<sup>d</sup> | n = XX<sup>d</sup> |
| *Regular* |  | XX/XX (%) | XX/XX (%) | XX/XX (%) | XX/XX (%) | XX/XX (%) | XX/XX (%) |
| *Irregular* |  | XX/XX (%) | XX/XX (%) | XX/XX (%) | XX/XX (%) | XX/XX (%) | XX/XX (%) |
| *Not Available<sup>e</sup>* | NA | XX/XX (%) | XX/XX (%) | XX/XX (%) | XX/XX (%) | XX/XX (%) | XX/XX (%) |
| *Missing* |  | XX/XX (%) | XX/XX (%) | XX/XX (%) | XX/XX (%) | XX/XX (%) | XX/XX (%) |

a= Patients who are alive, not withdrawn consent, continuing trial medication, who had in-hospital follow-up visit and completed medication form
b= Patients who are alive, not withdrawn, did not permanently discontinue trial medication till the previous follow-up visit and completed the medication form
c= Incident cases of permanent discontinuation at different visits.
d= Compliance as per treating physician (not applicable for patients have permanently discontinued the trial medication, and who had telephonic follow-up)
e= Patient who had telephonic follow-up

**Table 5.** Reasons of permanent discontinuation.

|  | 6 months | 12 months | 18 months | 24 months | 30 months | 36 months |
| --- | --- | --- | --- | --- | --- | --- |
| Permanently discontinued trial medication | XX | XX | XX | XX | XX | XX |
| Reasons (n) |  |  |  |  |  |  |
| *Valve surgery* | XX | XX | XX | XX | XX | XX |
| *Patient discontinued by themselves* | XX | XX | XX | XX | XX | XX |
| *Consulting physician discontinued* | XX | XX | XX | XX | XX | XX |
| *Patient is consulting other hospital* | XX | XX | XX | XX | XX | XX |
| *Suspected Digoxin toxicity* | XX | XX | XX | XX | XX | XX |
| *Moved to another city* | XX | XX | XX | XX | XX | XX |
| *Open label Digoxin* | XX | XX | XX | XX | XX | XX |

\*\*Note: Total of XX participants (XX%) have permanently discontinued the trial medication at different follow-up visits

**Table 6.**
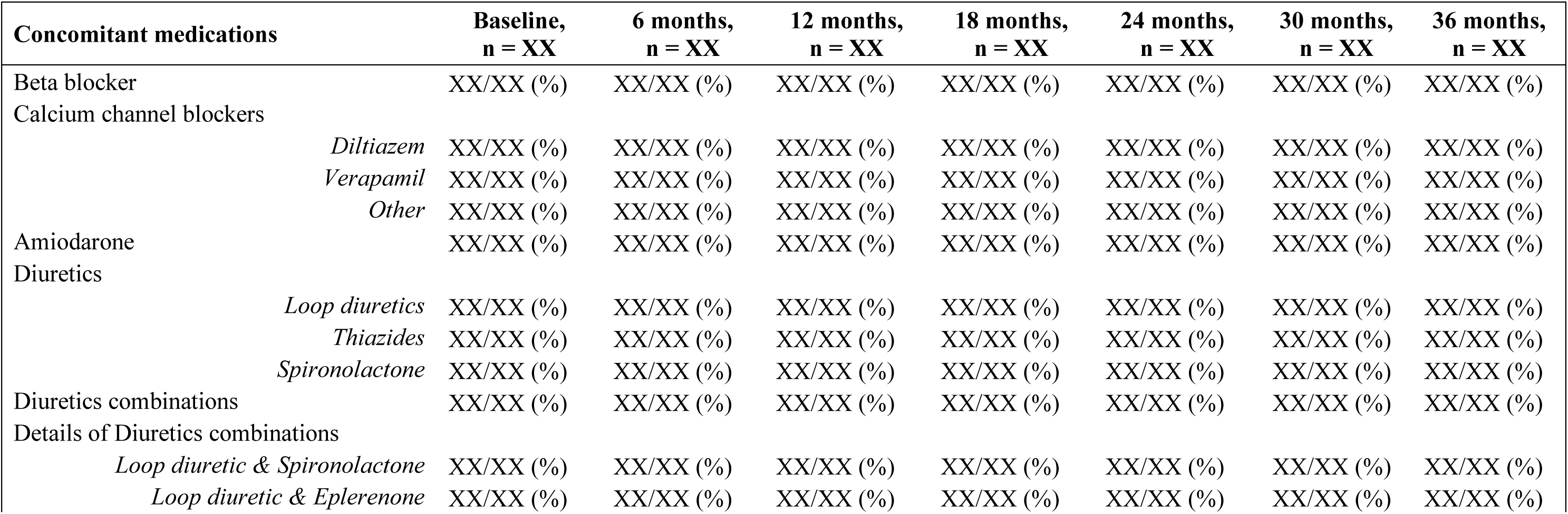
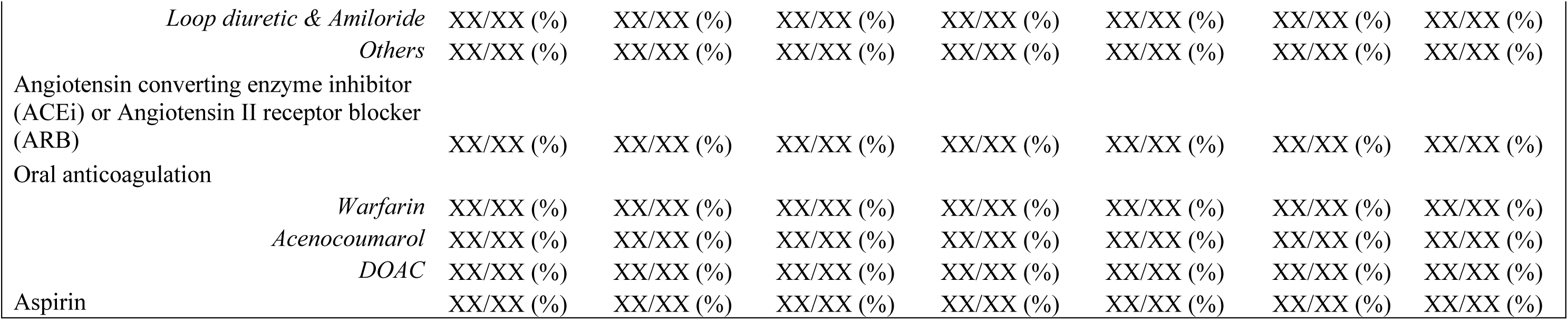

**Table 7.** Secondary prophylaxis.

|  | Baseline,<br>n = XX | 6 months,<br>n = XX | 12 months,<br>n = XX | 18 months,<br>n = XX | 24 months,<br>n = XX | 30 months,<br>n = XX | 36 months,<br>n = XX |
| --- | --- | --- | --- | --- | --- | --- | --- |
| None<sup>a</sup> | XX/XX | XX/XX | XX/XX | XX/XX | XX/XX | XX/XX | XX/XX |
| Penicillin IM<sup>b</sup> | XX/XX | XX/XX | XX/XX | XX/XX | XX/XX | XX/XX | XX/XX |
| Penicillin Oral<sup>b</sup> | XX/XX | XX/XX | XX/XX | XX/XX | XX/XX | XX/XX | XX/XX |
| Recently discontinued<sup>c</sup> | - | XX/XX | XX/XX | XX/XX | XX/XX | XX/XX | XX/XX |
| Compliance<sup>d</sup> |  |  |  |  |  |  |  |
| Regular | NA | XX/XX | XX/XX | XX/XX | XX/XX | XX/XX | XX/XX |
| Irregular |  | XX/XX | XX/XX | XX/XX | XX/XX | XX/XX | XX/XX |

a= Number of participants who were not on penicillin prophylaxis at baseline and during any of the follow ups. The denominator for the follow-up visits are those who were not on prophylaxis previously and have completed 6/12/18/24-month visits respectively.
b= Number of participants for whom Penicillin (IM/Oral) is initiated at current visit and those who have completed 6/12/18/24-month visit
c= Number of participants who have recently discontinued from Penicillin, out of those who had their Penicillin initiated at last follow-up visit and have completed 6/12/18/24-month follow-up visit.
d= Number of participants found taking Penicillin regularly/irregularly at current follow-up visit, out of those for whom Penicillin was initiated at their previous visit and have completed 6/12/18/24-month follow-up visit.

**Table 8.** Protocol deviation.

|  | **Digoxin** | **Placebo** |
| --- | --- | --- |
| Randomized ineligible patients | XX/XX (%) | XX/XX (%) |
| Discontinuation of medication<br>without advice of physician | XX/XX (%) | XX/XX (%) |

**Table 9.**
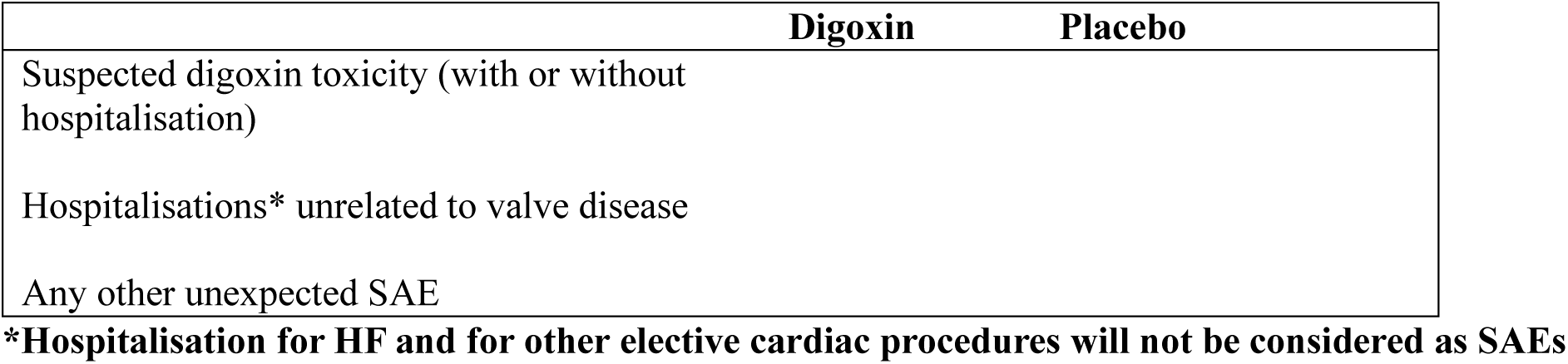
Serious adverse events.

**Table 10.**
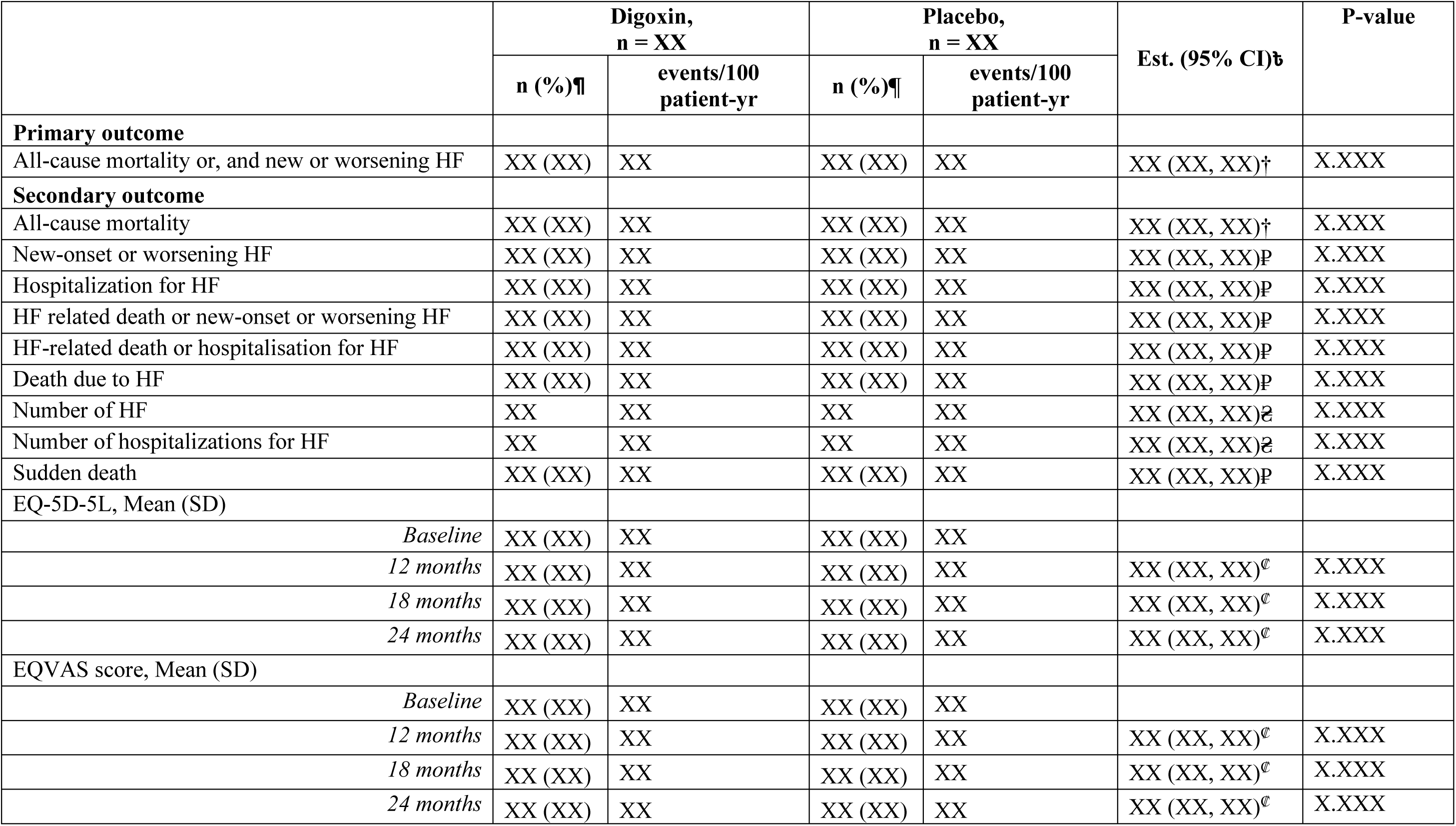
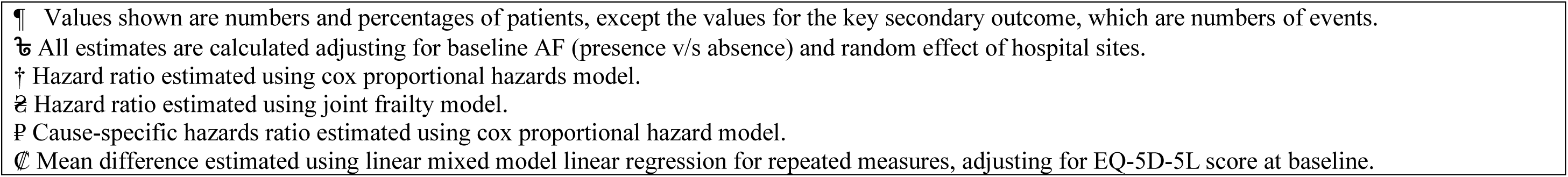
Main analysis of outcomes.

**Table 11.**

| Subgroup analysis |  | Digoxin v/s Placebo<br>HR (95% CI)† | P-value |
| --- | --- | --- | --- |
| **Primary outcome:** All-cause death or, and new or worsening HF, n/N |  |  |  |
| AF at baseline |  |  | XX |
|  | *Presence* | XX (XX, XX) |  |
|  | *Absence* | XX (XX, XX) |  |
| Digoxin therapy at the time of enrolment |  |  | XX |
|  | *Yes* | XX (XX, XX) |  |
|  | *No* | XX (XX, XX) |  |
| Sex |  |  | XX |
|  | *Male* | XX (XX, XX) |  |
|  | *Female* | XX (XX, XX) |  |

† = Hazard ratio estimated using cox proportional hazard model adjusting for AF (presence v/s absence) at baseline and random effect of hospital sites. For subgroup analysis, main effect for subgroup (if not already included) and a treatment-by-subgroup interaction term are included.

**Table 12.**
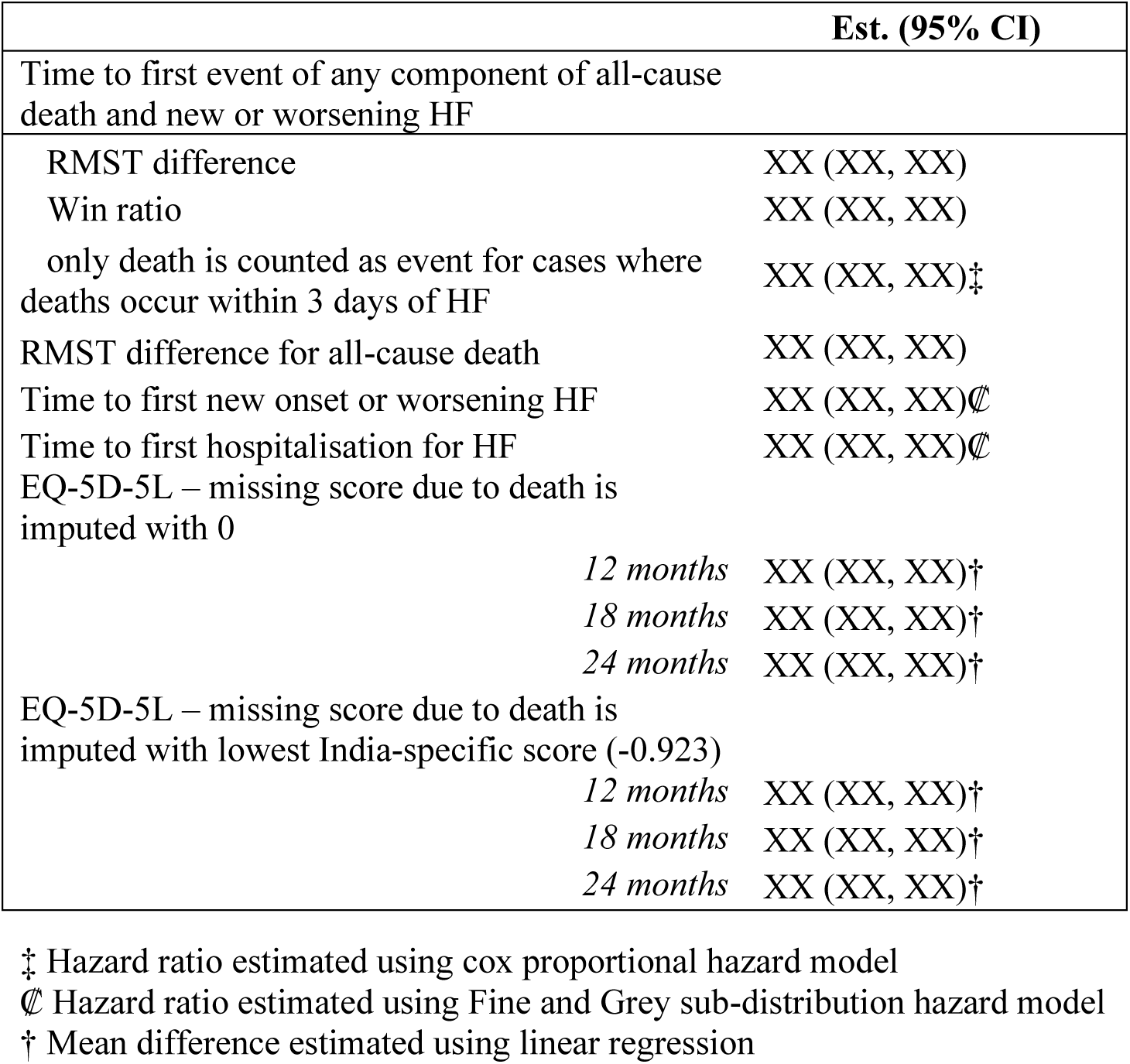
Sensitivity analysis for primary and secondary outcomes.

Figures

1. CONSORT diagram.
2. Cumulative incidence plot for primary outcome and the components.
3. Box plot of EQ-5D-5L scores at baseline and scores at baseline and 12, 18 and 24 months; and domain-wise bar charts
4. Distribution of wins and ties at each level of the hierarchy of the primary composite endpoint.

---

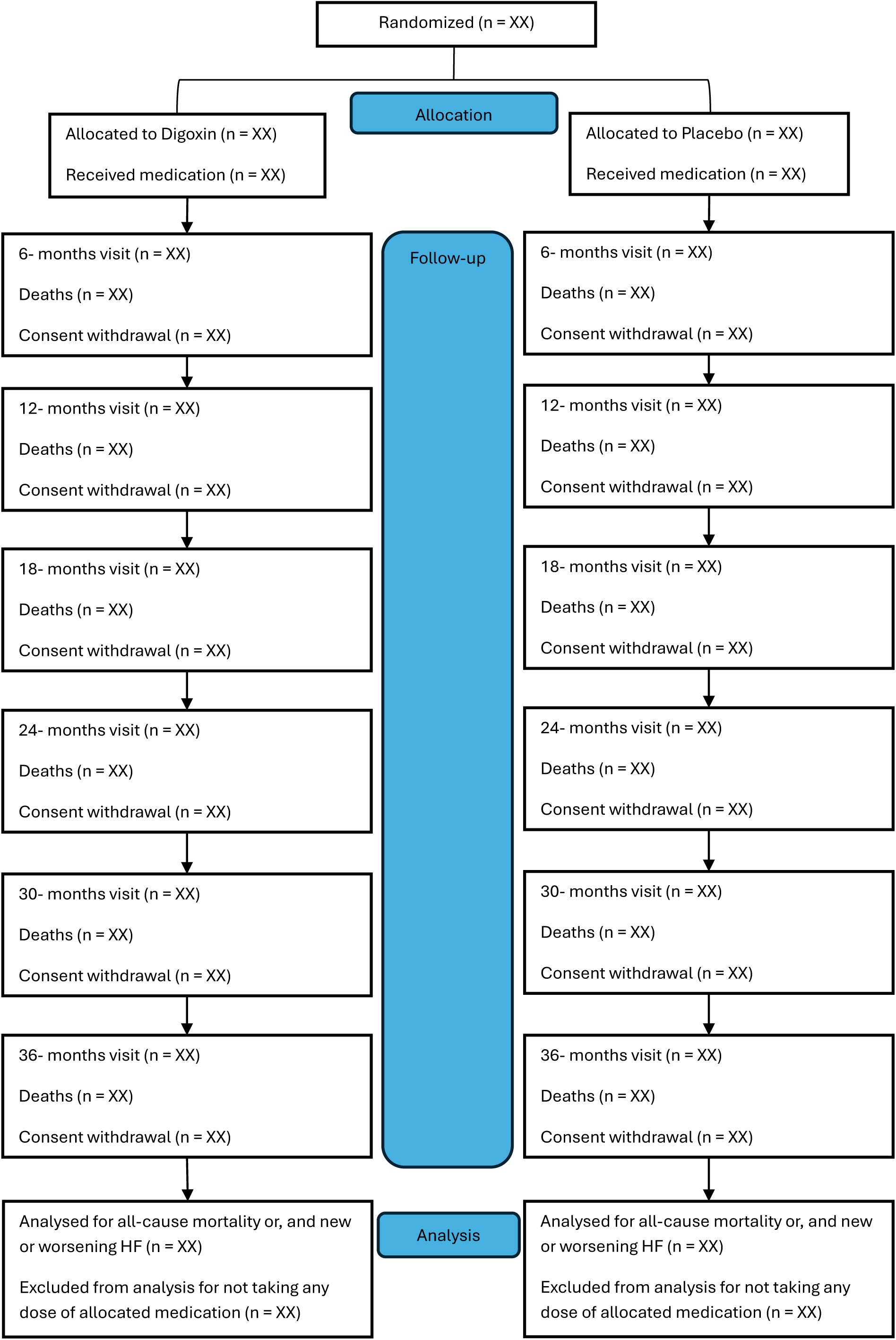

